# Global, Regional, and National Burden of Non-Rheumatic Calcified Aortic Valve Disease from 1990 to 2021 and Projections to 2050

**DOI:** 10.1101/2025.02.05.25321722

**Authors:** Zhenglong Wang, Hao Li, Hongwei Wei, Ming Chen, Xiaobo Liu

**Affiliations:** School of Clinical Medicine, Shandong Second Medical University, Weifang 261000, China; School of Clinical Medicine, Qingdao University, Qingdao 266000, China; Department of Cardiovascular Surgery, Affiliated Hospital of Shandong Second Medical University, Weifang 261035, China

**Author notes:** Corresponding author. Xiaobo Liu. Zhenglong Wang, Hao Li contributed equally to this article.

**Keywords:** non-rheumatic calcific aortic valve disease, Global burden of disease, Socio-demographic index, cardiology, epidemiology

## Abstract

**Background:** non-rheumatic calcific aortic valve disease (nrCAVD) is a valvular disease characterized by progressive calcification of the aortic valve that can lead to progressive left heart failure and death. This study reports on the global burden of CAVD, utilizing all available data and using the Global Burden of Disease (GBD) study methodology to understand the epidemiology of this widely prevalent disease.

**Methods:** Prior to the current effort, the burden of CAVD was included in GBD as a non-specific contributor to “valvular heart disease” burden. In this study, CAVD was distinguished as its own cause of death and disability in GBD, producing comparable and consistent estimates of CAVD burden. We used epidemiological and vital registry data to estimate the non-fatal and fatal burden of CAVD in 204 countries and 21 territories from 1990 to 2021 using standard GBD modelling approaches.

**Results:** In 2021, there were an estimated 13,320,896.13 (95% uncertainty interval [UI] 11,422,539.18 to 15,249,410.95) prevalent cases of CAVD globally. Of these, 6,018,666.77 (5,139,855.53 to 6,897,468.51) were in females (45%) and 7,302,229.37 (6,260,246.93 to 8,340,870.42) in males (55%). The age-standardised prevalence was 158.35 cases per 100 000 population (95% UI 135.92 – 181.00). Prevalence increased with age such that the highest prevalence was among individuals aged 90–94 years. In 2021, there were 142,205.00 例 (120,674.91 – 155,574.74) attributed to PAH globally, with an age-standardised mortality rate of 1.83 deaths from CAVD per 100 000 population (1.54 – 2.00). The burden of disease appears to be improving over time.

## 1. Introduction

Calcific aortic valve disease (CAVD) is a common heart valve disease characterized by progressive calcification of the aortic valve and is particularly prevalent in developed countries^[1, 2]^. Its main pathological feature is characterized by the deposition of calcium phosphate nodules in the fibrous matrix of the aortic valve leaflets, which gradually leads to valve dysfunction and stenosis^[3]^. In recent years, the incidence and prevalence of CAVD has increased significantly with the increasing trend of global aging. Data from 2013 showed that approximately 4.9 million older adults in Europe and 2.7 million in North America have been diagnosed with aortic stenosis, and the number of affected individuals is expected to increase globally from 2.5 million in 2000 to 4.5 million in 2030^[4, 5]^. Patients with severe aortic stenosis who are not treated with aortic valve replacement (AVR) have a 5-year survival rate of only 15%^[6]^.

CAVD is not only one of the leading causes of cardiovascular-related deaths in the elderly, but also a major public health challenge for healthcare systems worldwide. As the burden of disease increases, the direct and indirect healthcare costs associated with CAVD continue to climb, especially in developed countries^[7]^. There are no drugs that can effectively reverse or slow the progression of CAVD, and most patients eventually require transcatheter or surgical aortic valve replacement. However, there is no effective treatment other than replacement surgery to slow the natural course of CAVD ^[8, 9]^. The high morbidity and mortality of CAVD not only poses a serious threat to patients’ health, but also constitutes a heavy socio-economic burden on the global health care system^[10]^.

Previous studies have explored the global epidemiological characteristics of CAVD using Global Burden of Disease (GBD) data. The GBD 2017 analysis showed that the age-standardized incidence of calcific aortic valve disease remained essentially unchanged between 1990 and 2017, but that mortality rates and disability-adjusted life years (DALYs) increased significantly. This increase is largely attributable to population growth and aging trends. The prevalence, mortality, and overall burden of CAVD varied significantly geographically during this period, with high-income countries having the highest age-standardized DALY rates^[11]^. The recent GBD 2019 study further assessed the epidemiologic burden of CAVD at the global, regional, and national levels between 1990 and 2019, and found a 138% increase in the number of associated deaths globally, with particularly significant increases in DALYs, especially in countries with high sociodemographic indices (SDIs)^[12]^.

No study has yet conducted a comprehensive analysis of the global burden of CAVD using the latest GBD 2021 data. This study systematically assesses the global and regional distribution of CAVD, its temporal trends, and its impact on health based on the latest GBD 2021 data. By analyzing trends in morbidity, mortality, and disability-adjusted life years (DALYs), this study aims to clarify the public health significance of CAVD and provide a scientific basis for developing early screening, intervention, and treatment strategies.

## 2. Methods

### Data source

The migraine data of CAVD analyzed in this study are derived from the GBD 2021, which offers a comprehensive evaluation of health loss linked to 376 diseases, injuries, and impairments, along with 88 risk factors, across 204 nations and territories, employing the latest epidemiological data and refined standardized methods. The GBD database employs sophisticated methods to address missing data and adjust for confounding factors. Details about the study design and methods of GBD studies have been extensively described in existing GBD literature. Further information on the location, disease, and risk hierarchies used for GBD 2021 can be found on the Global Health Data Exchange (GHDx). The University of Washington’s Institutional Review Board approved the use of de-identified data for the GBD study and waived the requirement for informed consent.

### General statistical methods

Estimates were generated for each GBD age group, location, year, and sex. Uncertainty was estimated by sampling from the posterior distributions at each step in the modelling process and is reported as 95% uncertainty intervals (UIs), with lower and upper bounds calculated as the 2.5% and 97.5% quantile. Three significant figures were used when reporting estimates. While this is consistent with convention in GBD, this means that component estimates presented in the manuscript (eg, males and females) cannot always be added to generate the reported total. We used the direct method with the GBD standard global population to produce age-standardised estimates. The GBD standard population is based on the population structure of all countries with populations greater than 5 million people. In a two-step process, the proportion of the location-specific population in each age group is calculated and then these age-specific proportions are averaged across all locations. Code used for GBD estimation is publicly available online.

### Joinpoint regression analysis

The joinpoint regression model is a collection of linear statistical models that were used to evaluate the trends in disease burdens attributable to CAVD across time. This model’s calculating approach is to estimate the changing rule of illness rates using the least square method, avoiding the non-objectivity of typical trend analyses based on linear trends. Calculating the square sum of the residual error between the estimated and actual values yields the turning point of the shifting trend. Joinpoint (version 5.3.0.0; National Cancer Institute, Rockville, MD, USA) was used to create this model. We also calculated the average annual percentage change (AAPC) and investigated if the fluctuation trend in different parts was statistically significant by comparing the AAPC to 0. A statistically significant P-value was less than 0.05.

### Age-period-cohort analysis

The age-period-cohort model examines the effect of age, period and cohort on health outcomes. Age effect refers to the risk of outcomes at different ages. Period effect refers to the effect of temporal changes on outcomes across all age groups. Cohort effect refers to the changes in outcomes among participants with the same birth cohorts. The log-linear regression model is expressed as: log(Yi) = μ + α ∗ age_i_ + β ∗ period_i_ + γ ∗ cohort_i_ + ε, where Yi is the CAVD prevalence or deaths rate, α, β and γ are the coefficients of age, period and cohort, respectively, μ is the intercept and ε is the residual of model. The intrinsic estimator (IE) method integrated into age-period-cohort model was used to get the net effects for three dimensions.

### Decomposition analysis and frontier analysis

Decomposition analysis was used to visualize the role of three factors (aging, demographic, and epidemiological) that drive changes in DALYs between 1990 and 2021. Epidemiologic change refers to potential age- and population-adjusted mortality and morbidity. We applied frontier analyses to further assess the relationship between CAVD burden and sociodemographic development. To generate a nonlinear frontier that implies the lowest achievable burden based on developmental status. We used nonparametric data envelopment analysis and referred to detailed descriptions of previous studies. The distance between a country’s observed disability-adjusted life-years (DALY) ratio and its boundary (defined as the effective variance) represents the unrealized health gains that exist based on the current level of development in the country or region.

### Measurement health inequalities

Total DALYs and age-age standardized DALY rates were extracted for inequality analysis. As per the recommendations of the World Health Organization, two standard measures, namely Slope Index of Inequality (SII) and Concentration Index, were used to assess both absolute and relative income-related inequalities between countries. The SII represents the slope of the regression line that correlates the country-level age-standardised DALY rate (ASDR) associated with CAVD with the weighted ranking of each country. To adjust for variations in burden levels, the SII is divided by the global ASDR, yielding the Relative Index of Inequality (RII). The CI is utilized to assess the relative disparity in the burden of CAVD among countries by fitting the Lorenz concentration curve based on cumulative DALYs and cumulative population. The CI is a numerical integration of the area under the curve, ranging from − 1 to 1. A negative CI value indicates a higher concentration of CAVD burden among populations residing in countries with lower SDI.

### Ethics

The institutional review board granted an exemption for this study, as it utilized publicly accessible data that contained no confidential or personally identifiable patient information.

## 3. Results

### Descriptive analysis

In 2021, there were 13 320 896.13 prevalent cases (95% UI 11 422 539.18 – 15 249 410.95) of CAVD around the world. The age-standardised prevalence of CAVD was 158.35 cases per 100 000 population (95% UI 135.92 – 181.00). Estimates of CAVD prevalence appeared to be relatively stable over time, with an age-standardised prevalence of CAVD in 1990 of 129.75 per 100 000 population (106.01 – 152.98; Table 1). CAVD was more prevalent among males, with 156.31 cases of CAVD per 100 000 males (128.67 – 183.78) versus 128.88 cases of CAVD per 100 000 females (110.13 – 147.63) in 2021.Of the 13 320 896.13 individuals with prevalent CAVD, 7 302 229.37 (6 260 246.93 – 8 340 870.42) were male (55%) and 6 018 666.77 (5 139 855.53 – 6 897 468.51) were female (45%). Figure 1 shows the age-standardised prevalence and distribution of CAVD by sex and age globally. There was a different CAVD prevalence worldwide, with largely higher prevalence in western Europe, central Europe, and high-income Asia North America, and a lower prevalence in east Asia, south Asia, southeast Asia, and all over Africa. Age-standardised prevalence by region ranged from 11.08 cases per 100 000 population (8.99 – 13.41) in Western Sub-Saharan Africa to 425.32 cases per 100 000 population (375.41 – 477.97) in western Europe. Prevalence of CAVD increased with age, with the highest prevalence observed in individuals aged 90–94 years.

**Table. 1.** Prevalence of CAVD in 1990 and 2021 for both sexes and all locations.

| Location | 1990 |  | 2021 |  |
| --- | --- | --- | --- | --- |
|  | Cases (95% UI) | Age-standardized<br>prevalence per 100 000<br>population (95% UI) | Cases (95% UI) | Age-standardized<br>prevalence per 100 000<br>population (95% UI) |
| Global | 4,686,909.62<br>(3,874,992.36 to 5,539,482.79) | 129.75<br>(106.01 to 152.98) | 13,320,896.13<br>(11,422,539.18 to 15,249,410.95) | 158.35<br>(135.92 to 181.00) |
| Female | 2,193,824.73<br>(1,785,712.61 to 2,614,908.40) | 108.21<br>(87.83 to 127.91) | 6,018,666.77<br>(5,139,855.53 to 6,897,468.51) | 128.88<br>(110.13 to 147.63) |
| Male | 2,493,084.89<br>(2,060,652.51 to 2,942,214.55) | 156.31<br>(128.67 to 183.78) | 7,302,229.37<br>(6,260,246.93 to 8,340,870.42) | 193.24<br>(166.56 to 220.38) |
| High SDI | 3,032,967.42<br>(2,503,938.60 to 3,578,606.31) | 267.39 (221.28 to 314.24) | 7,860,601.13<br>(6,819,445.28 to 8,892,888.68) | 349.64 (303.58 to 395.77) |
| High-middle SDI | 1,153,180.99<br>(949,563.34 to 1,377,066.42) | 121.46 (99.65 to 145.53) | 3,440,879.34<br>(2,962,648.59 to 3,931,536.98) | 172.89 (149.52 to 197.05) |
| Middle SDI | 321,383.52<br>(259,294.94 to 390,497.74) | 33.93 (27.06 to 40.86) | 1,417,738.74<br>(1,134,289.02 to 1,688,923.53) | 54.49 (43.43 to 64.98) |
| Low-middle SDI | 141,850.08 (113,799.76 to 172,848.31) | 25.36 (20.48 to 30.85) | 496,397.06 (404,011.70 to 589,829.17) | 36.62 (29.46 to 43.57) |
| Low SDI | 30,541.60 (24,426.31 to 37,928.11) | 14.16 (11.28 to 17.30) | 85,816.14 (68,867.81 to 104,073.56) | 17.64 (14.26 to 21.19) |
| Central Asia | 37,213.16 (29,704.23 to 45,741.96) | 82.14 (65.58 to 100.76) | 113,425.99 (93,422.35 to 135,263.82) | 146.37 (119.97 to 173.94) |
| Central Europe | 295,016.25 (242,496.22 to 353,469.99) | 197.53 (163.07 to 236.16) | 866,860.01 (755,597.05 to 995,396.94) | 379.57 (330.44 to 437.56) |
| Eastern Europe | 377,604.70 (300,011.56 to 463,741.53) | 136.91 (109.03 to 167.66) | 949,894.85 (761,766.27 to 1,153,178.23) | 264.72 (213.79 to 319.46) |
| Australasia | 51,082.99 (42,840.73 to 59,259.85) | 214.44 (179.76 to 247.44) | 192,488.44 (165,456.40 to 223,129.60) | 332.99 (286.39 to 384.01) |
| High-income Asia Pacific | 482,345.31<br>(385,464.86 to 591,334.60) | 244.46<br>(194.31 to 297.30) | 1,447,553.37<br>(1,198,036.50 to 1,699,764.57) | 284.22<br>(238.29 to 333.61) |
| High-income North America | 1,196,674.50<br>(943,508.39 to 1,460,579.62) | 325.59<br>(256.18 to 394.83) | 2,772,453.82<br>(2,360,163.14 to 3,202,756.36) | 403.07<br>(344.88 to 462.72) |
| Southern Latin America | 72,336.25 (60,572.88 to 83,630.69) | 159.61 (133.14 to 184.61) | 234,828.04 (202,977.28 to 273,602.42) | 262.02 (226.21 to 304.68) |
| Western Europe | 1,602,827.79<br>(1,382,936.40 to 1,854,393.70) | 264.82<br>(228.72 to 306.36) | 4,306,127.72<br>(3,789,648.99 to 4,809,887.14) | 425.32<br>(375.41 to 477.97) |
| Andean Latin America | 14,076.47 (11,665.03 to 17,291.07) | 71.52 (58.70 to 88.00) | 77,587.91 (66,209.70 to 89,727.71) | 133.19 (113.31 to 154.29) |
| Caribbean | 23,568.52 (19,825.85 to 27,881.94) | 91.96 (77.05 to 108.80) | 72,780.51 (62,508.09 to 84,939.19) | 134.62 (115.73 to 157.17) |
| Central Latin America | 82,893.68 (67,918.34 to 99,369.27) | 105.40 (85.42 to 127.23) | 395,444.67 (324,616.09 to 470,756.28) | 160.32 (131.28 to 191.12) |
| Tropical Latin America | 93,905.59 (76,978.30 to 113,891.78) | 105.97 (86.08 to 129.68) | 355,803.79 (284,980.97 to 430,929.73) | 139.74 (111.90 to 169.21) |
| North Africa and Middle East | 63,862.15 (52,184.02 to 76,625.74) | 38.28 (31.34 to 45.70) | 246,279.61 (205,246.82 to 291,455.07) | 55.12 (46.06 to 64.89) |
| South Asia | 96,317.75 (75,479.46 to 120,424.36) | 18.31 (14.40 to 22.93) | 349,852.67 (276,806.33 to 429,247.39) | 24.88 (19.64 to 30.42) |
| East Asia | 137,958.02 (104,340.22 to 174,295.82) | 17.53 (13.34 to 21.90) | 719,161.48 (554,740.08 to 884,832.44) | 33.15 (25.50 to 40.55) |
| Oceania | 494.60 (390.27 to 614.34) | 20.10 (15.94 to 24.23) | 1,670.62 (1,321.76 to 2,032.32) | 27.09 (21.54 to 32.45) |
| Southeast Asia | 33,921.71 (25,960.04 to 42,234.73) | 15.06 (11.64 to 18.35) | 150,953.26 (115,913.59 to 183,273.86) | 25.31 (19.72 to 30.59) |
| Central Sub-Saharan Africa | 2,664.67 (2,106.43 to 3,335.26) | 12.49 (9.93 to 15.42) | 7,684.17 (6,108.63 to 9,323.96) | 14.67 (11.85 to 17.89) |
| Eastern Sub-Saharan Africa | 8,446.05 (6,750.53 to 10,487.17) | 11.63 (9.22 to 14.28) | 22,929.15 (18,357.29 to 27,771.09) | 13.96 (11.03 to 17.01) |
| Southern Sub-Saharan Africa | 5,111.82 (4,072.84 to 6,265.97) | 18.87 (14.96 to 23.03) | 13,442.15 (10,821.30 to 16,419.98) | 23.77 (19.01 to 28.80) |
| Western Sub-Saharan Africa | 8,587.63 (6,868.29 to 10,575.58) | 9.18 (7.36 to 11.19) | 23,673.89 (19,211.64 to 28,588.83) | 11.08 (8.99 to 13.41) |

**Figure. 1:**
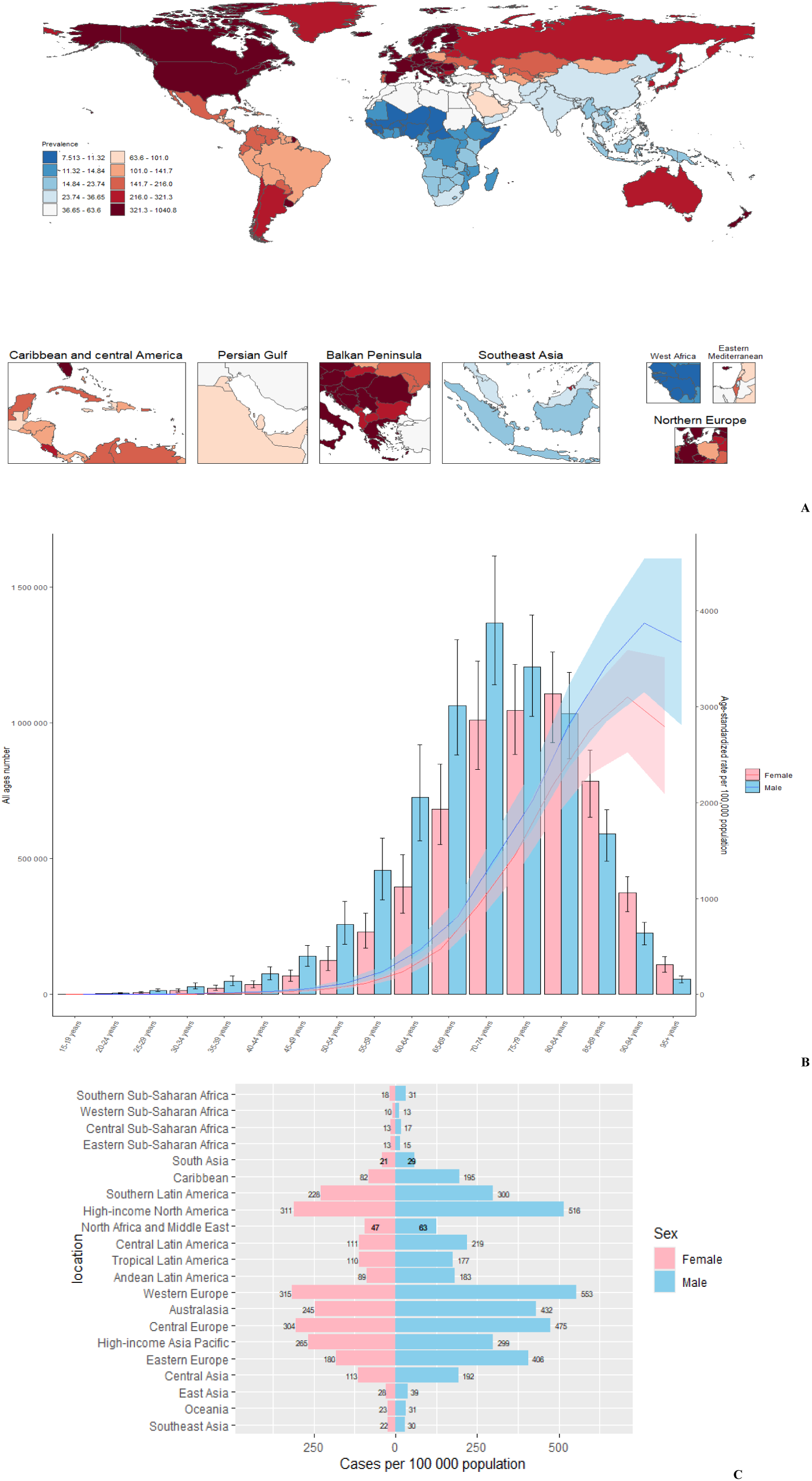
CAVD prevalence (A) Age-standardised prevalence of CAVD by region in 2021 (both sexes). (B) Global age-specific prevalence of CAVD in 2021; Bar graphs are all-age numbers; line graphs are age-standardized rates. (C) Age-standardised prevalence of CAVD by sex and region in 2021.

In 2021, there were 142 205.00 deaths (95% UI 120 674.91 – 155 574.74) attributed to CAVD globally. The age-standardised mortality rate attributed to CAVD (cause-specific mortality) in 2021 was 1.83 deaths per 100 000 population (95% UI 1.54 – 2.00). Cause-specific mortality improved slowly over time as the 2021 estimate was lower than the estimated rate in 1990 of 1.92 deaths from CAVD per 100 000 population (1.73 – 2.06), a decrease of 4.7% (Table 2).

**Table. 2.** Mortality of CAVD in 1990 and 2021 for both sexes and all locations.

| Location | 1990 |  | 2021 |  |
| --- | --- | --- | --- | --- |
|  | Cases (95% UI) | Age-standardized<br>mortality per 100 000<br>population (95% UI) | Cases (95% UI) | Age-standardized<br>mortality per 100 000<br>population (95% UI) |
| Global | 57,931.98 (52,886.03 to 61,949.99) | 1.92 (1.73 to 2.06) | 142,205.00 (120,674.91 to 155,574.74) | 1.83 (1.54 to 2.00) |
| Female | 32,636.78 (28,827.69 to 35,762.87) | 1.84 (1.60 to 2.01) | 82,555.58 (66,341.27 to 92,742.90) | 1.73 (1.40 to 1.95) |
| Male | 25,295.20 (23,583.35 to 27,103.98) | 1.94 (1.80 to 2.07) | 59,649.42 (53,124.75 to 63,372.10) | 1.89 (1.66 to 2.02) |
| High SDI | 41,209.37 (37,108.99 to 43,310.25) | 3.78 (3.37 to 3.99) | 92,578.75 (74,403.69 to 102,070.93) | 3.43 (2.81 to 3.76) |
| High-middle SDI | 7,642.42 (7,067.04 to 8,132.14) | 0.96 (0.88 to 1.03) | 23,645.61 (20,185.73 to 25,780.96) | 1.27 (1.08 to 1.39) |
| Middle SDI | 4,177.85 (3,663.96 to 4,807.25) | 0.49 (0.42 to 0.57) | 11,639.63 (10,228.46 to 13,303.28) | 0.49 (0.43 to 0.57) |
| Low-middle SDI | 3,346.07 (2,384.47 to 4,455.53) | 0.67 (0.47 to 0.89) | 10,222.60 (8,152.25 to 12,208.75) | 0.84 (0.67 to 1.01) |
| Low SDI | 1,508.23 (894.92 to 2,161.11) | 0.86 (0.50 to 1.21) | 3,951.15 (2,656.06 to 5,097.33) | 0.98 (0.64 to 1.31) |
| Central Asia | 33.04 (28.83 to 38.68) | 0.07 (0.06 to 0.09) | 205.01 (178.50 to 231.10) | 0.30 (0.26 to 0.34) |
| Central Europe | 1,040.39 (956.03 to 1,179.06) | 0.75 (0.69 to 0.86) | 6,366.20 (5,693.48 to 6,899.51) | 2.68 (2.40 to 2.91) |
| Eastern Europe | 351.95 (337.61 to 364.00) | 0.14 (0.13 to 0.14) | 2,230.04 (2,062.67 to 2,390.70) | 0.64 (0.59 to 0.69) |
| Australasia | 807.33 (729.99 to 861.18) | 3.63 (3.25 to 3.87) | 2,002.14 (1,661.94 to 2,221.40) | 3.09 (2.59 to 3.42) |
| High-income Asia Pacific | 6,320.89 (5,644.60 to 6,687.27) | 3.74 (3.28 to 4.00) | 19,320.05 (13,988.05 to 22,319.09) | 2.46 (1.84 to 2.80) |
| High-income North America | 14,039.48 (12,326.54 to 14,903.97) | 3.83 (3.36 to 4.07) | 27,281.95 (22,212.01 to 29,882.54) | 3.60 (2.97 to 3.93) |
| Southern Latin America | 1,653.64 (1,497.42 to 1,797.09) | 4.01 (3.61 to 4.37) | 2,764.67 (2,448.75 to 3,010.92) | 3.01 (2.67 to 3.27) |
| Western Europe | 23,489.59 (21,550.22 to 24,746.33) | 3.97 (3.62 to 4.18) | 53,518.41 (43,825.48 to 58,591.63) | 4.23 (3.53 to 4.60) |
| Andean Latin America | 130.85 (106.05 to 151.26) | 0.64 (0.52 to 0.74) | 356.34 (293.13 to 435.58) | 0.61 (0.50 to 0.74) |
| Caribbean | 248.36 (220.79 to 270.58) | 0.99 (0.89 to 1.08) | 545.25 (476.33 to 613.95) | 1.01 (0.88 to 1.14) |
| Central Latin America | 700.32 (674.48 to 717.89) | 0.88 (0.84 to 0.90) | 2,225.73 (1,984.86 to 2,480.35) | 0.91 (0.82 to 1.02) |
| Tropical Latin America | 1,595.55 (1,524.46 to 1,643.37) | 1.88 (1.76 to 1.96) | 3,837.36 (3,444.06 to 3,444.06) | 1.55 (1.39 to 1.66) |
| North Africa and Middle East | 1,418.08 (961.99 to 1,873.83) | 0.92 (0.60 to 1.20) | 3,275.61 (2,414.14 to 4,031.97) | 0.81 (0.61 to 0.99) |
| South Asia | 3,175.52 (2,106.11 to 4,454.30) | 0.72 (0.47 to 1.00) | 11,191.70 (8,667.81 to 13,803.93) | 0.92 (0.70 to 1.13) |
| East Asia | 812.94 (550.51 to 1,178.48) | 0.13 (0.09 to 0.18) | 1,757.12 (1,421.98 to 2,248.05) | 0.10 (0.08 to 0.12) |
| Oceania | 24.29 (17.59 to 35.65) | 1.08 (0.83 to 1.48) | 54.21 (39.93 to 76.69) | 0.84 (0.64 to 1.20) |
| Southeast Asia | 363.33 (234.86 to 665.69) | 0.19 (0.12 to 0.36) | 1,242.75 (941.53 to 1,950.71) | 0.25 (0.19 to 0.39) |
| Central Sub-Saharan Africa | 206.75 (129.91 to 294.14) | 1.27 (0.77 to 1.79) | 569.19 (351.99 to 857.16) | 1.45 (0.88 to 2.27) |
| Eastern Sub-Saharan Africa | 572.46 (351.19 to 804.02) | 0.98 (0.57 to 1.34) | 1,378.76 (899.05 to 1,885.38) | 1.02 (0.66 to 1.44) |
| Southern Sub-Saharan Africa | 226.55 (177.96 to 276.36) | 0.99 (0.75 to 1.20) | 525.42 (425.52 to 626.49) | 1.23 (0.98 to 1.45) |
| Western Sub-Saharan Africa | 720.66 (447.47 to 960.96) | 1.06 (0.64 to 1.48) | 1,557.10 (966.13 to 2,404.16) | 1.04 (0.65 to 1.63) |

Because a higher prevalence of CAVD in males, cause-specific mortality in the population was similar as prevalence, with 1.94 deaths from CAVD per 100 000 males (1.80 – 2.07) and 1.84 deaths from CAVD per 100 000 females (1.60 – 2.01) in 2021. Figure 2 shows cause-specific mortality per 100 000 population and the distribution by sex and age globally. Mortality attributed to CAVD was different worldwide, with largely higher cause-specific mortality in Western Europe, Australasia, and High-income North America, and lower mortality in east Asia, south Asia, Central Asia and Andean Latin America. Age-standardised cause-specific mortality by region ranged from 0·10 deaths from CAVD per 100 000 population (0.08 – 0.12) in East Asia to 4.23 deaths from CAVD per 100 000 population (3.53 – 4.60) in Western Europe.

**Figure. 2:**
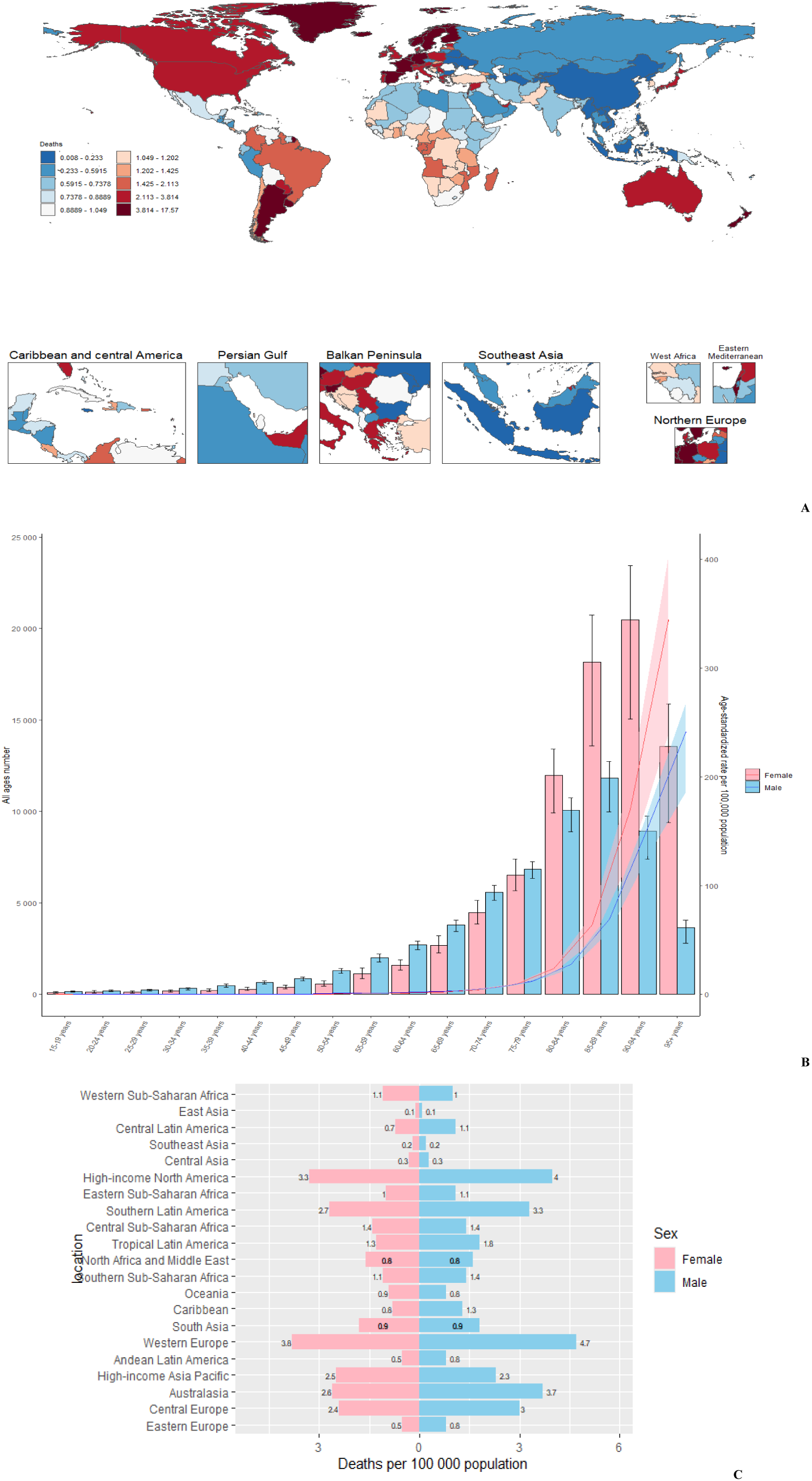
CAVD mortality (A) Age-standardised mortality attributed to CAVD by region in 2021 (both sexes). (B) Total global deaths from CAVD by age group in 2021; Bar graphs are all-age numbers; line graphs are age-standardized rates. (C) Age-standardised mortality attributed to CAVD by sex and region in 2021.

Figure 3 depicts the trends in the sex-specific all-age number and age-standardized rates of CAVD incidence and mortality in Global from 1990 to 2021. The age-standardized rates of the four indicators are relatively stable, resulting in a steady increase of all-age cases by calendar year. Interestingly, the age-standardized rates for all four indicators were higher for men than for women, but the all-age cases of deaths and DALYs were higher for women than for men (Figure 3A,3B).

**Figure 3.**
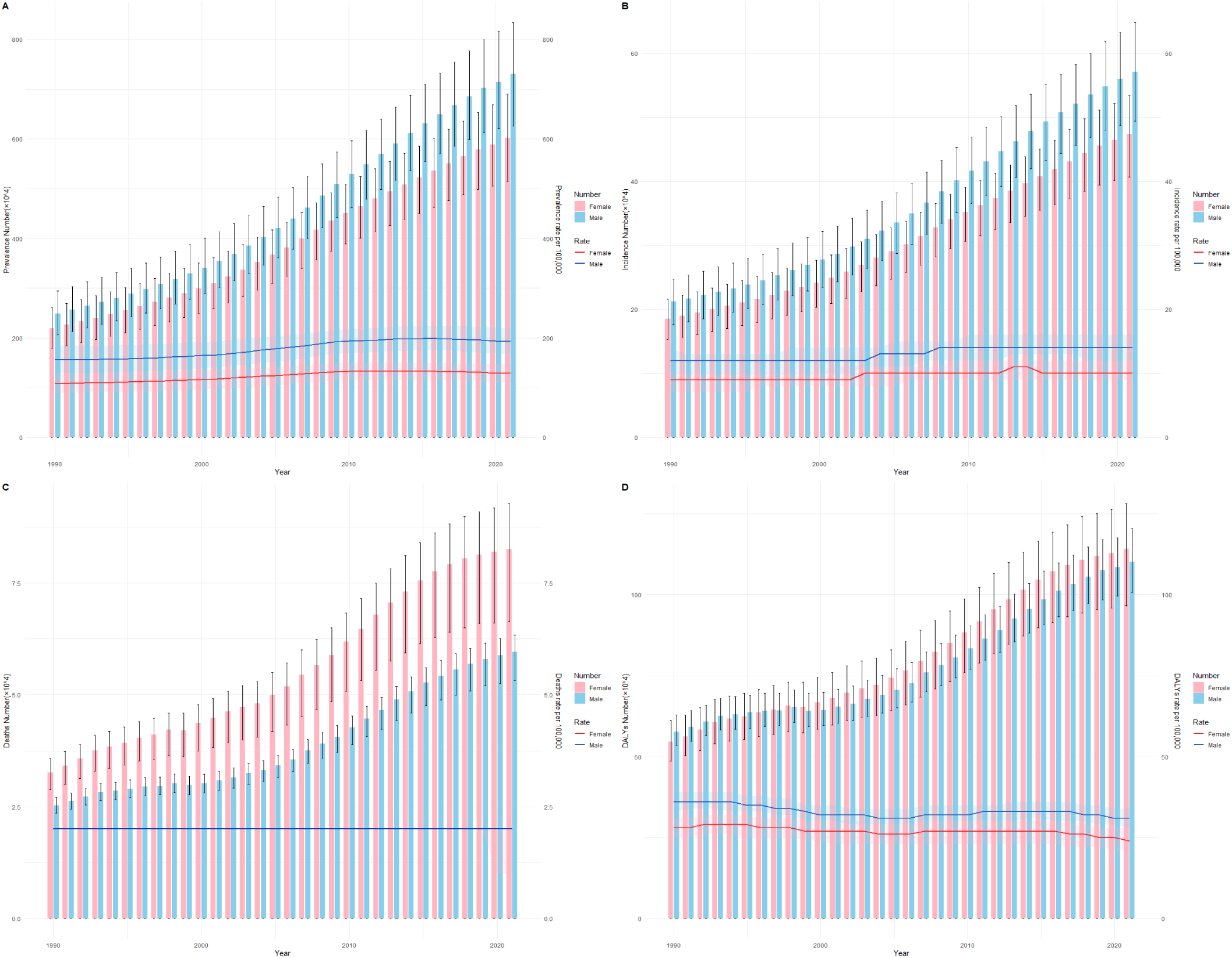
Trends in the all-age cases and age-standardized rates of CAVD by sex from 1990 to 2021. (A) prevalence. (B) Incidence. (C)Deaths. (D)DALYs

### Joinpoint regression analysis

Joinpoint regression analyses of the age-standardized prevalence rates for CAVD in global from 1990 to 2021 are shown in Figure 4. We found the disease prevalence trend to significantly increase from 2001 to 2010 in male (APC=+1.77 (2001–2010), 95% CI: 1.71, 1.83) (Figure 4A), female (APC=+1.54 (2001–2009), 95% CI: 1.49, 1.60) (Figure 4B) and both sexes (APC=+1.68 (2001–2009), 95% CI: 1.62, 1.74) populations (Figure 4C). Since 2010, the prevalence trend has moderated in both sexes. However, it is still rising year on year, and the incidence is on a similar upward trend. Albeit the age-standardized mortality rate significantly decreased in both males (1993–2000 APC=−1.35, 2015–2021 APC=−1.42) and females (1993–2004 APC=−0.52, 2015–2021 APC=−2.10), it increased in both males (1990– 1993 APC=1.67, 2004–2015 APC=1.09) and females (1990–1993 APC=1.98, 2004–2015 APC=0.62) (Figures 5A,5B). The cases of both sexes are similar as females (Figure 5C).

**Figure. 4.**
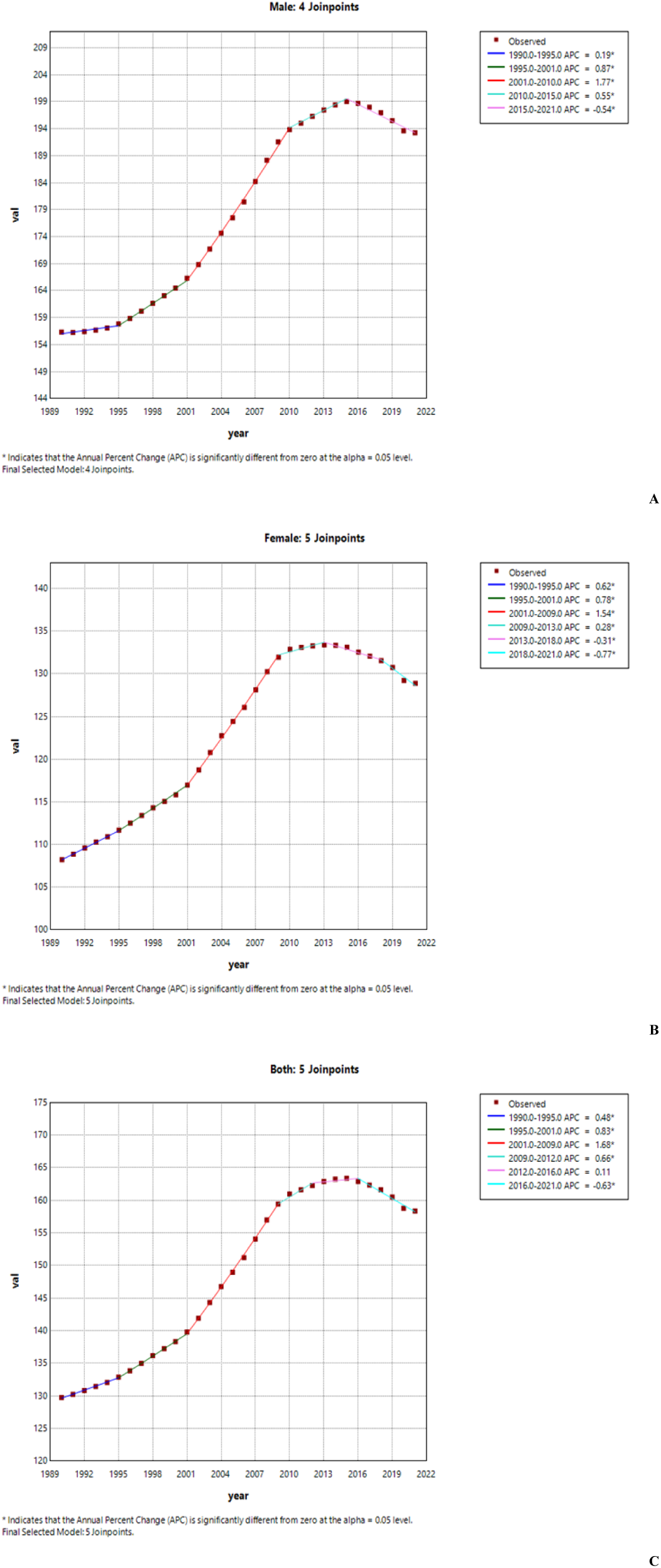
Joinpoint regression analysis of the sex-specific age-standardized prevalence rate for CAVD in global from 1990 to 2021. (A) Age-standardized prevalence rate for males. (B) Age-standardized prevalence rate for females. (C) Age-standardized prevalence rate for both sexes.

**Figure. 5.**
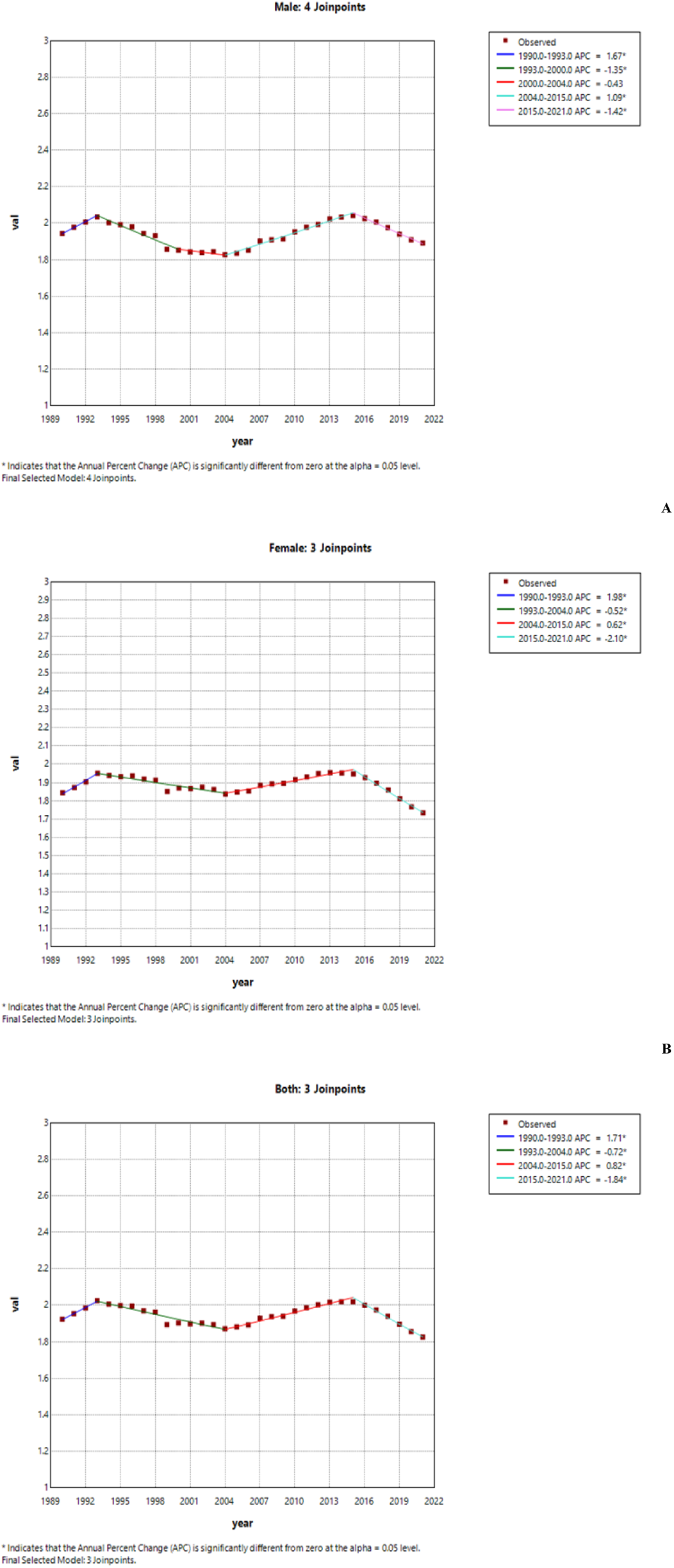
Joinpoint regression analysis of the sex-specific age-standardized mortality rate for CAVD in global from 1990 to 2021. (A) Age-standardized mortality rate for males. (B) Age-standardized mortality rate for females. (C) Age-standardized prevalence rate for both sexes.

Table 3 shows the AAPCs in CAVD prevalence, and mortality rates over three decades. Age-standardized prevalence and mortality rates for CAVD in global increased by 0.64 (95% CI: 0.59, 0.70) and −0.16 (95% CI: −0.30, −0.03), respectively, from 1990 to 2021. Surprisingly, females had a lower AAPC of prevalence, and mortality rates than males (Table 3).

**Table. 3.** AAPC, average annual percent change presented for full period; APC, annual percent change; CI, confidence interval.

| Gender | ASPR |  |  | ASMR |  |  |
| --- | --- | --- | --- | --- | --- | --- |
|  | Period | APC (95% CI) | AAPC (95% CI) | Period | APC (95% CI) | AAPC (95% CI) |
| **Both** | 1990-1995 | 0.48 (0.36-0.60) | 0.64 (0.59-0.70) | 1990-1993 | 1.71 (0.84-2.59) | -0.16 (-0.30--0.03) |
|  | 1995-2001 | 0.83 (0.72-0.94) |  | 1993-2004 | -0.72 (-0.87--0.57) |  |
|  | 2001-2009 | 1.68 (1.62-1.74) |  | 2004-2015 | 0.82 (0.64-1.00) |  |
|  | 2009-2012 | 0.66 (0.28-1.04) |  | 2015-2021 | -1.84 (-2.25--1.43) |  |
|  | 2012-2016 | 0.11 (-0.08--0.30) |  |  |  |  |
|  | 2016-2021 | -0.63 (-0.72--0.55) |  |  |  |  |
| **Female** | 1990-1995 | 0.62 (0.51-0.73) | 0.56 (0.52-0.60) | 1990-1993 | 1.98 (1.12-2.85) | -0.19 (-0.31--0.06) |
|  | 1995-2001 | 0.78 (0.68-0.89) |  | 1993-2004 | -0.52 (-0.66--0.38) |  |
|  | 2001-2009 | 1.54 (1.49-1.60) |  | 2004-2015 | 0.62 (0.45-0.79) |  |
|  | 2009-2013 | 0.28 (0.10-0.46) |  | 2015-2021 | -2.10 (-2.48--1.71) |  |
|  | 2013-2018 | -0.31 (-0.42--0.20) |  |  |  |  |
|  | 2018-2021 | -0.77 (-0.96--0.58) |  |  |  |  |
| **Male** | 1990-1995 | 0.19 (0.04-0.33) | 0.69 (0.65-0.74) | 1990-1993 | 1.67 (0.81-2.54) | -0.09 (-0.27-0.08) |
|  | 1995-2001 | 0.87 (0.73-1.01) |  | 1993-2000 | -1.35 (-1.64--1.05) |  |
|  | 2001-2010 | 1.77 (1.71-1.83) |  | 2000-2004 | -0.43 (-1.38-0.52) |  |
|  | 2010-2015 | 0.55 (0.40-0.69) |  | 2004-2015 | 1.09 (0.93-1.26) |  |
|  | 2015-2021 | -0.54 (-0.62--0.45) |  | 2015-2021 | -1.42 (-1.80--1.04) |  |

### The effects of age, period, and cohort on prevalence and mortality rates

Figure 6 illustrated the age-period-cohort effect of CAVD prevalence and mortality rate. The CAVD prevalence increased from the ages of 55–59 years, peaking at the ages of 90–94 years (Figure 6A). The period-based trends of CAVD prevalence were relatively steady in the younger groups and trended upward over the period in the 80–95+ age groups (Figure 6B). The RR value increased from 0.95 (95% CI 0.94–0.96) in 1994 to 1.08 (95% CI 1.07–1.09) in 2014. The birth cohort of each age group showed that the CAVD prevalence in the early period was lower than that in the later period (Figure 6C). The cohort risk was significantly lower in the early birth cohort (RR cohort (1902–1907) = 0.61, 95% CI 0.58–0.64) and increased in the recent cohorts (RR cohort (2002–2007) = 1.18, 95% CI 0.69–2.03). In contrast, the increase in age-specific CAVD mortality rate was exponentially distributed. The increase of RR values was initially slow in the groups aged under 59 years old and then became exponential (RR age (75–79) = 9.68, 95% CI 9.49–9.87) (Figure 6D). The period-based mortality rate showed a clear downward trend until 2004 (RR period (2004) = 1.00, 95% CI 1.00–1.00) and leveled off between 2004 and 2009 (RR period (2009) = 1.00, 95% CI 0.99–1.01), but gradually increased between 2009 and 2014 (RR period (2014) = 1.03, 95% CI 1.01–1.04) and then decreased afterwards (RR period (2019) = 0.99, 95% CI 0.97–1.00) (Figure 6E). Similarly, the early birth cohort has a greater effect on CAVD mortality. The mortality rate peaked in 1937 (RR cohort (1922) = 1.42, 95% CI 1.39–1.45) and then decreased afterwards [RR period (1927) = 1.33, 95% CI 1.30–1.36]. Then the RR displayed a downward trend from 1.22 in the 1927–1932 birth cohort to 0.70 in the 1997–2002 birth cohort (Figure 6F). Notably, those borned between 1977 and 1982 had a slightly higher mortality rate.

**Figure. 6.**
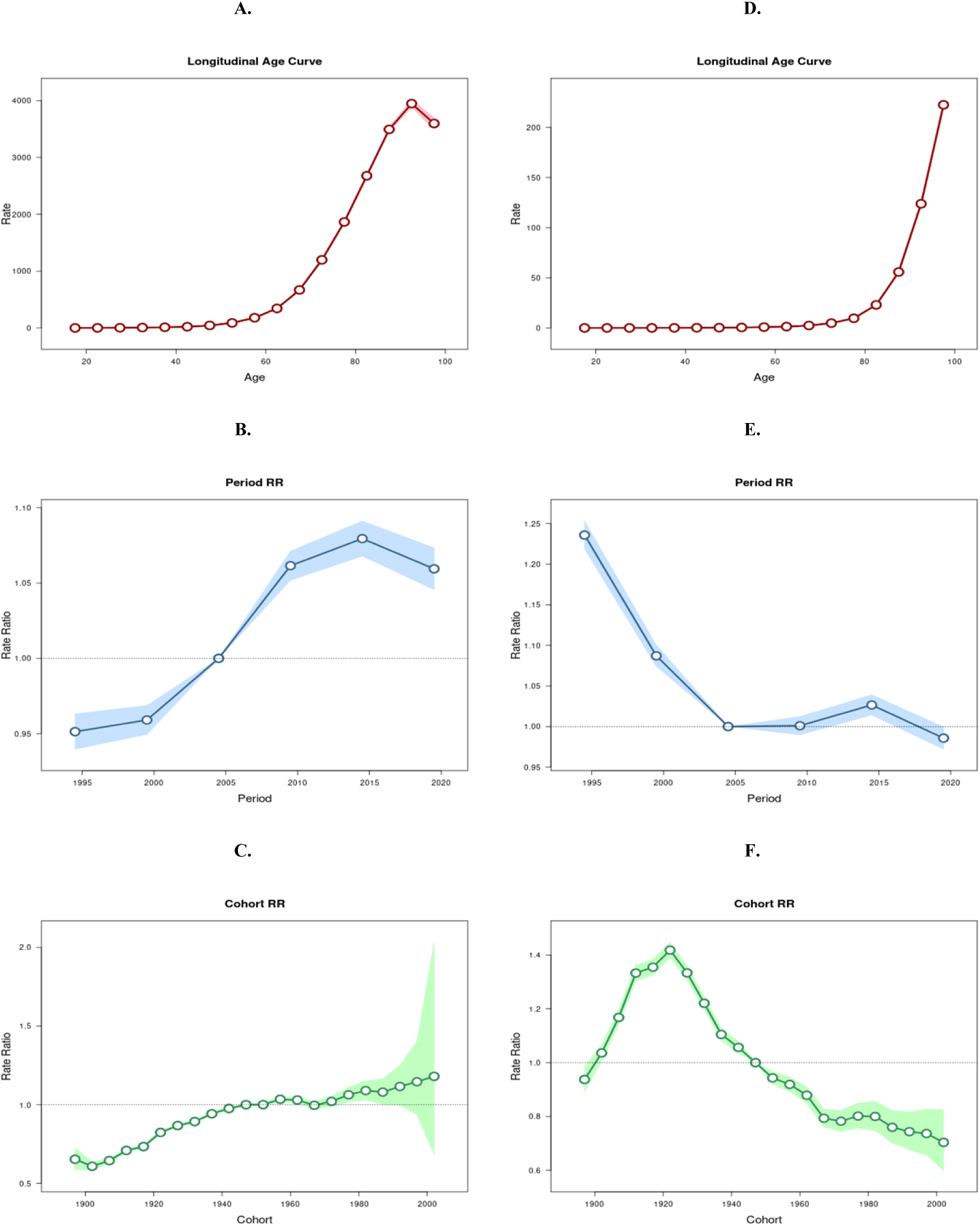
Age, period and cohort effects on CAVD prevalence and mortality rate in global during 1990–2021. (A–C) prevalence rate; (D–F) mortality rate.

**Figure. 7.**
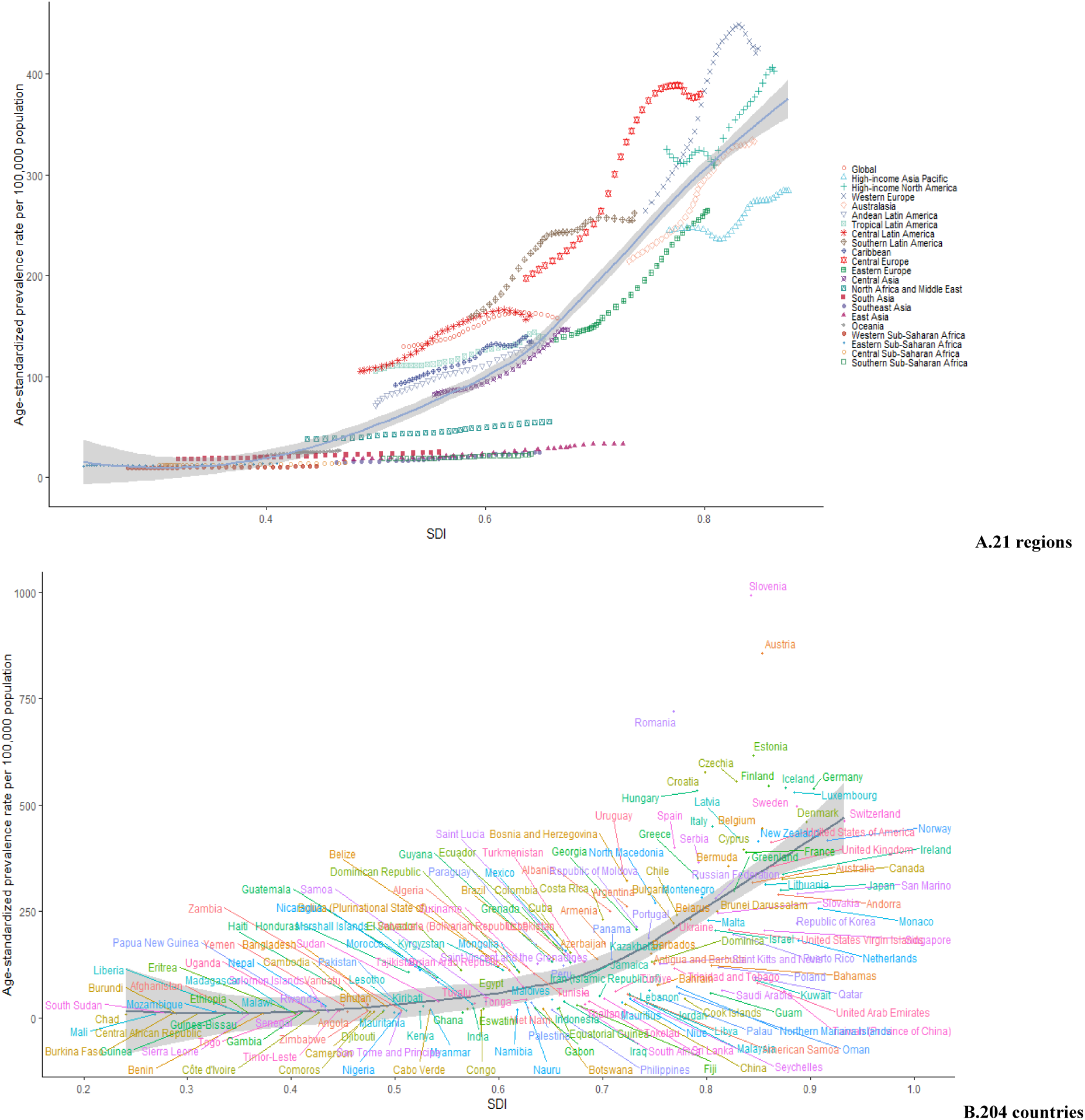
The associations between the SDI and prevalent rates per 100,000 population of migraine in CAVD across 21 GBD regions. CAVD = calcific aortic valve disease, SDI = Socio-Demographic Index, GBD = Global Burden of Disease

### The association between migraine burden and SDI

In 2021, there was a positive correlation between prevalence rates, incidence rates, and DALYs rates of migraine in CAVD and the SDI. With the improvement of the economy, the overall disease burden is on the rise. Over-all, the global burden of migraine is slightly higher than expected. Across 21 regions, the burden of migraine in CAVD remains relatively stable when the SDI value falls below 0.5. The burden of migraine reaches its peak when the SDI reaches 0.8. Regions such as Southern Latin America, Central Europe and Western Europe have a higher burden than expected, while regions such as Southern Sub-Saharan Africa, Southeast Asia, and East Asia have a lower burden than expected. In 204 countries, the burden of migraine in Slovenia, Romania, Austria, and Germany exceeds expectations, while in China, Monaco, Canada, and Japan, the burden is lower than anticipated (Figure7).

### Decomposition analysis of age-standardized DALYs rates

The past 30 years have seen a significant global increase in DALYs, with the largest increase occurring in middle SDI quintile regions (Figure 8). Aging and population growth accounted for 69.87% and 56.81%, of the worldwide increase in DALYs, respectively, with the most significant aging contribution occurring in the middle SDI quintile (60.87%) and most significant population growth occurring in the low SDI quintile (86.28%). The effect of epidemiological change on DALYs growth was negative (−26.69%) worldwide, and this effect was the most pronounced in the middle SDI quintile (−2.71%). This result may be due to the strong effects of aging and population growth. The effects of demography and epidemiology on DALYs differed across countries and regions

**Figure. 8.**
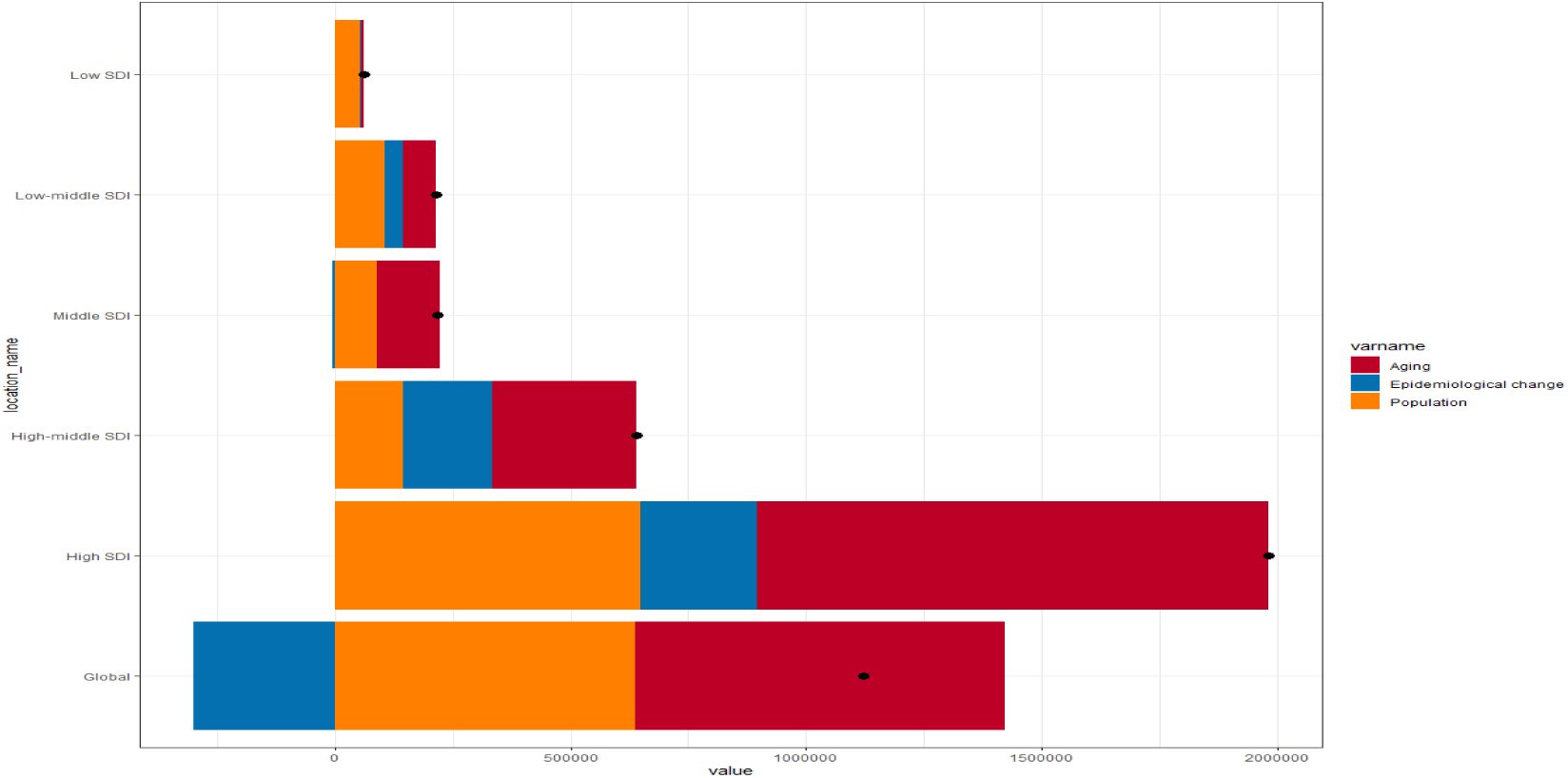
Changes in calcific aortic valve disease DALYs according to population-level determinants of population growth, aging, and epidemiological change from 1990 to 2019 at the global level and by SDI quintile. The black dot represents the overall value of change contributed by all 3 components. For each component, the magnitude of a positive value indicates a corresponding increase in calcific aortic valve disease DALYs attributed to the component; the magnitude of a negative value indicates a corresponding decrease in calcific aortic valve disease DALYs attributed to the related component.

### Frontier analysis of age-standardized DALYs rates

The unrealized health gains of countries or regions at different levels of development during the period 1990–2021 are shown in Figure 9A. As a supplement, figure 9B show the DALYs burden and the effective difference in countries or regions with different sociodemographic development levels in 2021. With sociodemographic development, effective difference generally increased to some extent, indicating that countries or regions with a higher SDI have greater burden improvement potential (Figure 9B).

**Figure. 9.**
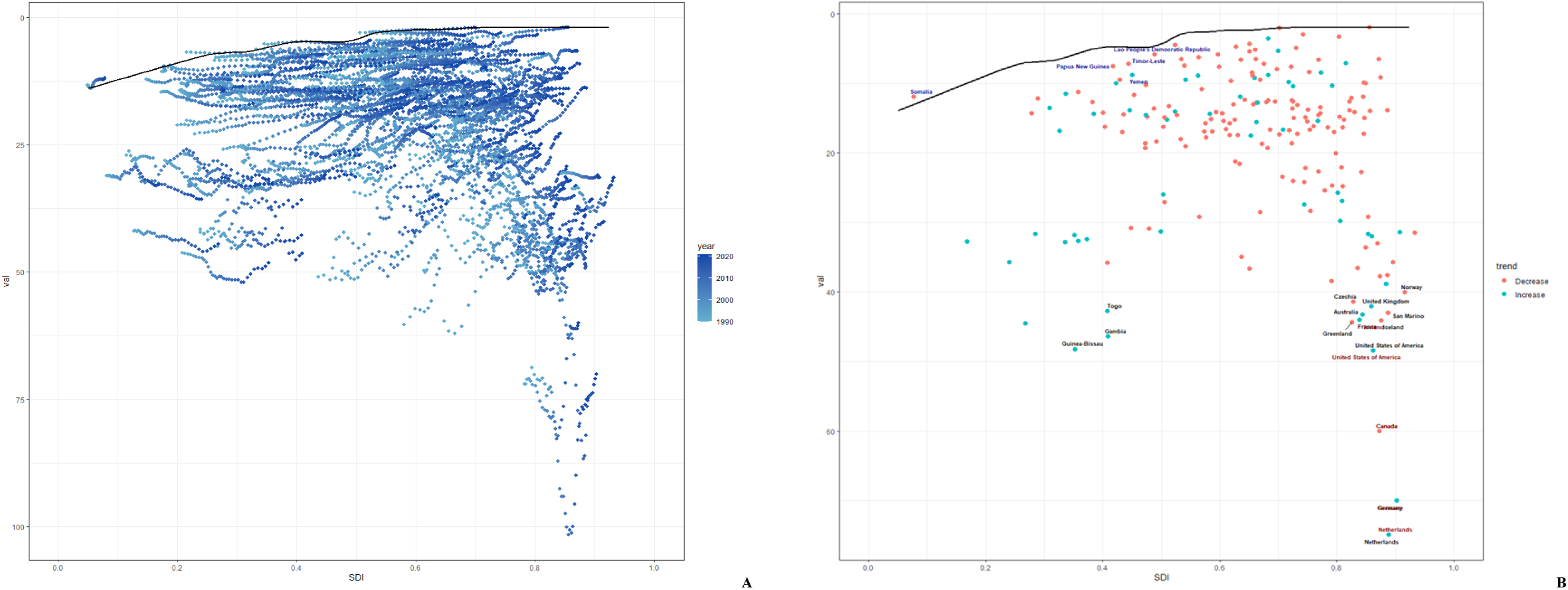
Frontier analysis based on SDI and age-standardized calcific aortic valve disease DALY rate in 2021. The frontier is delineated in solid black color; countries and territories are represented as dots. The top 15 countries with the largest effective difference (largest calcific aortic valve disease DALYs gap from the frontier) are labeled in black; examples of frontier countries with low SDI (< 0.5) and low effective difference are labeled in blue, and examples of countries and territories with high SDI (> 0.85) and relatively high effective difference for their level of development are labeled in red. Red dots indicate an increase in age-standardized calcific aortic valve disease DALYs rate from 1990 to 2021; blue dots indicate a decrease in age-standardized calcific aortic valve disease DALYs rate between 1990 and 2021.

### Cross-national HHD health inequality

In 1990 and 2021, the SII (per 100,000 population) for DALYs were −3.39517 (95% CI −38.0851 - 63.79712) and 1.391411 (95% CI −61.4884 - 85.18115) respectively, suggests a positive correlation between all-age DALY rates and the SDI index (Figure. 10). This slight increase suggests that inequality in the burden of all-age CAVDs between high-income and low-income countries increased during this period. Between 1990 and 2021, the concentration index for DALYs and Deaths has shown an upward trend. Although the inequality in the burden of CAVD has decreased regionally between poor and wealthy countries, inequality still persists. This indicates that while the wealth gap has narrowed in some regions, global inequality in CAVD remains a persistent issue (Figure. 11).

**Figure. 10.**
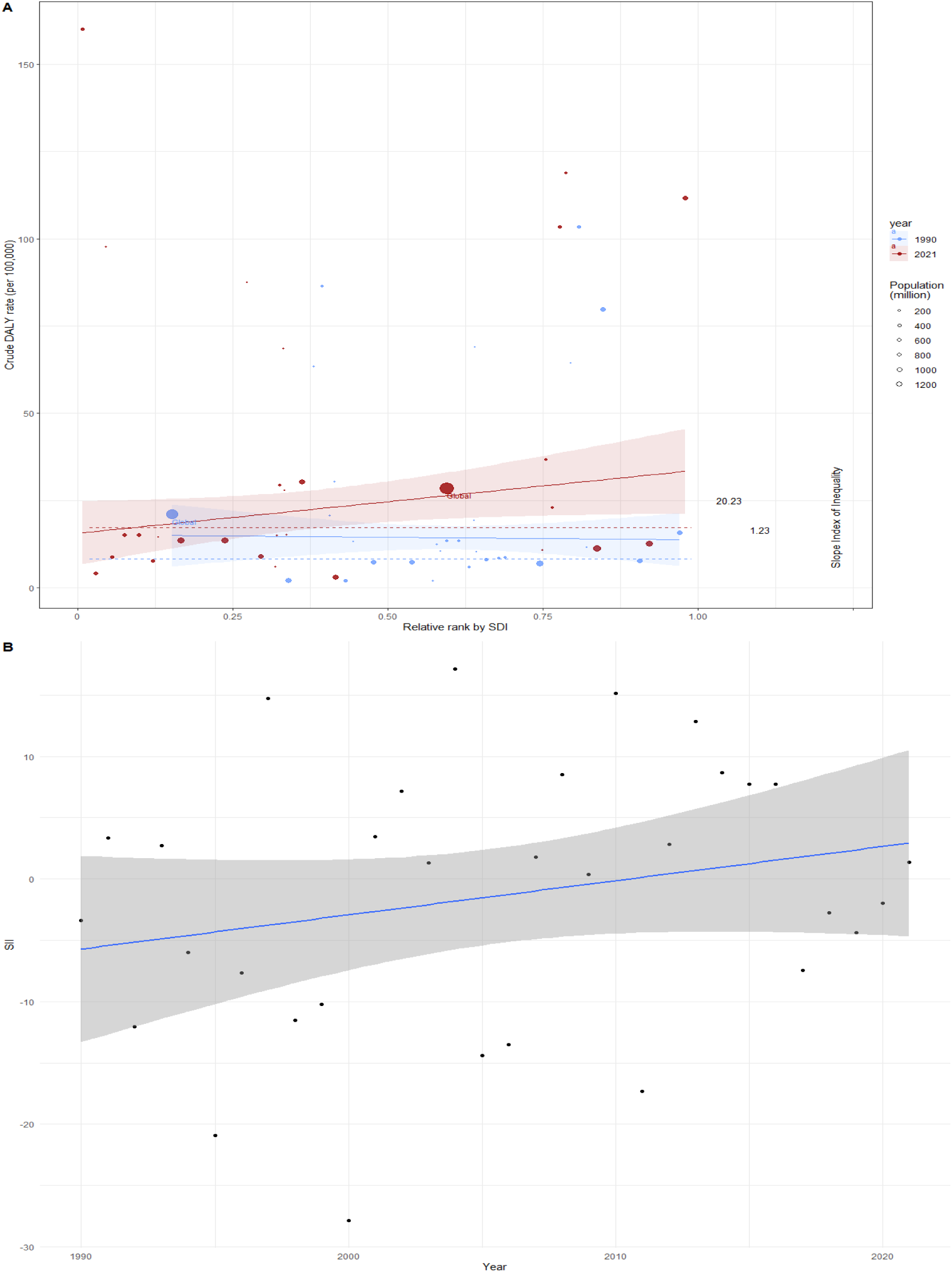
SII analysis. (A) Absolute income-related healthy inequality in CAVD burden, presented using regression lines, 1990 vs. 2021. (B) Trendline demonstrates the trend in SII from 1990 to 2021

**Figure. 11.**
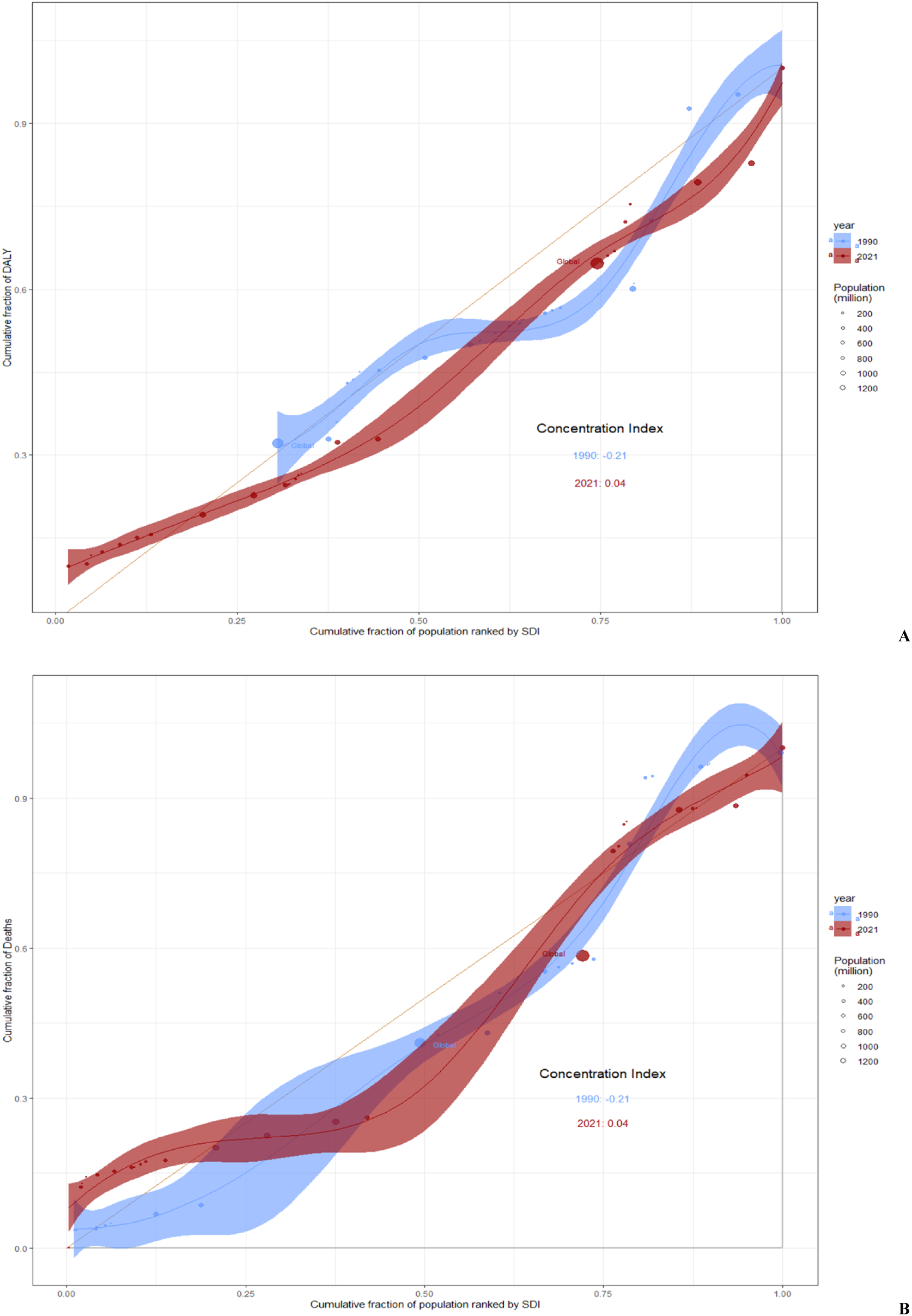
Concentration index analysis. (A) Relative income-related healthy inequality in CAVD burden, presented using concentration curves, 1990 vs. 2021. (B) Trendline demonstrates the trend in concentration index from 1990 to 2021

### Future forecasts of global burden of CAVD

The global burden of CAVD is projected to evolve significantly from 2021 to 2050, with varying trends across different measures. The ASPR of CAVD is expected to increase globally for both sexes. For females, the prevalence is expected to remain relatively stable, with a slight decrease from about 130 per 100,000 in 2021 to 103 per 100,000 in 2050. In contrast, males are projected to experience a more pronounced increase, from approximately 195 per 100,000 in 2021 to 159 per 100,000 in 2050, indicating a steeper downward trend (Figures 12A,12D). Compared with prevalence, the global incidence of CAVD also shows a slight downward trend for both sexes, with the ASIR of females projected to decrease from about 10 per 100,000 population in 2021 to approximately 7 per 100,000 by 2050. Similarly, the ASIR of males projected to decrease from about 14 per 100,000 population in 2021 to approximately 12 per 100,000 by 2050 (Figures 12B,12E). The age-standardized DALY rate for CAVD is expected to remain relatively stable for both sexes, with a slight decrease in females from 24.9 per 100,000 population in 2021 to approximately 18.9 per 100,000 by 2050. In contrast, males are projected to experience a more pronounced increase, from approximately 31.5 per 100,000 in 2021 to 24 per 100,000 in 2050 (Figures 12C,12F). Across all three measures, males consistently show higher rates compared to females, although the gap between males and females appears to narrow over time, particularly for prevalence.

**Figure. 12:**
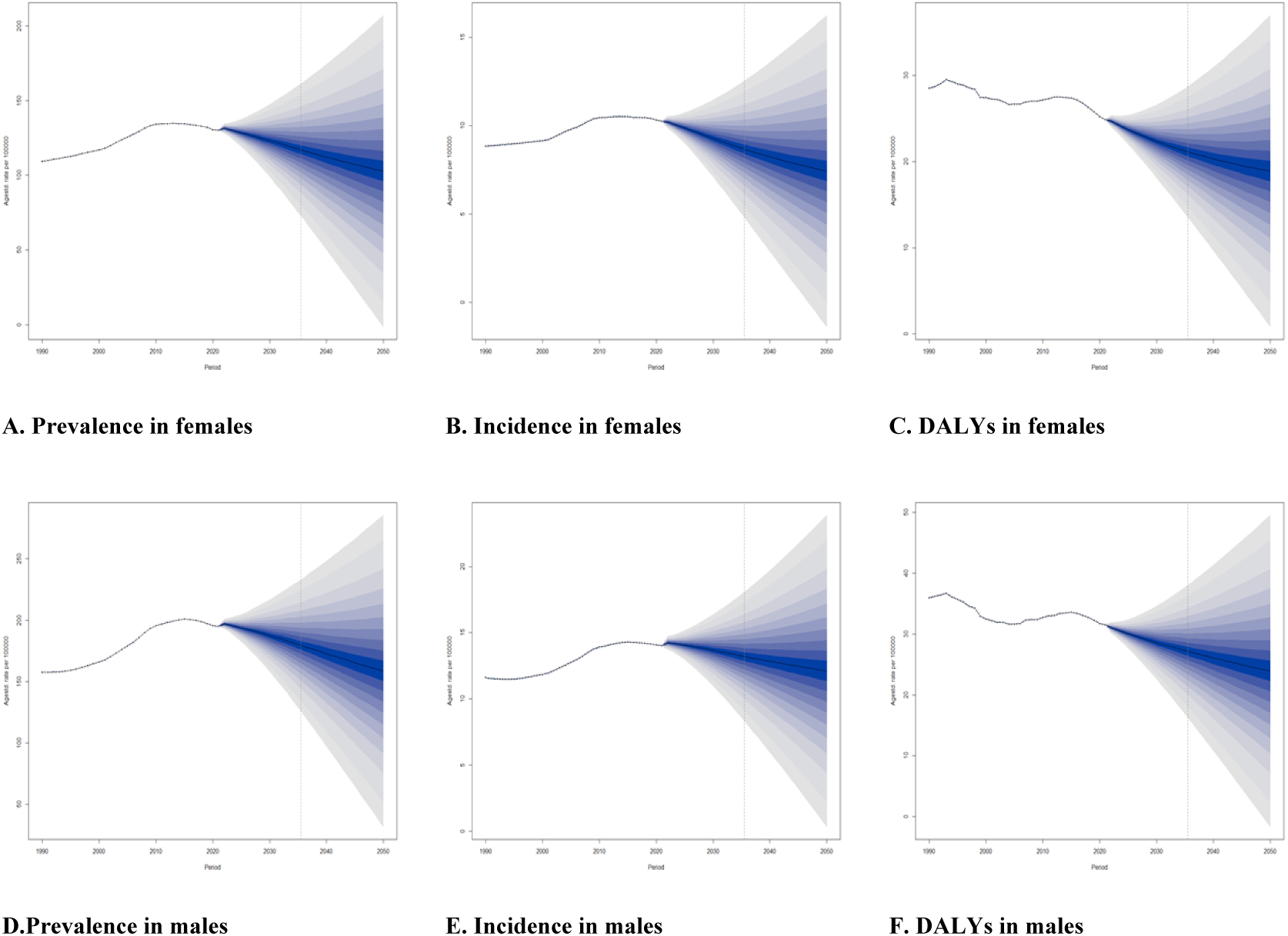
Future forecasts of global burden of CAVD.

## 4. Discussion

After decades of relative quiescence, the management of patients with aortic valve disease is again garnering interest due to the convergence of several factors^[13]^. Prevalence of calcific aortic valve disease is growing as the global population ages^[14, 15]^. Because of the relative commonness of CAVD, it imposes a considerable health burden worldwide^[16]^. In 2021, There were 158.35 prevalent cases of CAVD per 100 000 population, with a mortality rate of 1.83 deaths from CAVD per 100 000 population. However, there was modest variability across a relatively narrow range in both the prevalence and cause-specific mortality from CAVD worldwide. The burden from CAVD might be modestly improving over time. Previous studies, largely anchored to local or national registries, now the current GBD prevalence estimate of 158.35 prevalent cases of CAVD per 100 000 population using a more standardized and comprehensive dataset is similar to the mean of prevalence estimates in these previously reported studies, but substantially higher than estimates reported in previous reviews of global CAVD prevalence that relied on fewer data sources. We note that the ASIP and ASPR for CAVD have been gradually increasing since 1990, but seem to turn around in 2020 and will continue to decline for the next 30 years. Meanwhile, in our prediction model, ASIR and ASPR have been on a steady decline from 2021 to 2050. In contrast to the first two, DALY shows a fluctuating decline. Although the prevalence of CAVD remains high at present, programmatic improvements have mitigated patient prognosis to some extent by reducing the number of years of life lost (a key component of DALY).

Regional variations were observed in estimates of prevalence and mortality. In regions with high prevalence, the difference was approximately 55-fold higher compared with regions with low prevalence. Similarly, regions with high cause-specific mortality had an approximately 47-fold higher rate than those with lower mortality. These regional differences might reflect an uneven regional distribution of risk factors implicated in the pathogenesis of CAVD. For example, genetic pre-disposition^[17]^, dietary habits, frequency of exercise, and infections such as rheumatic fever vary by region and might drive local differences in CAVD prevalence^[16, 18]^. In addition, competing risks from infection, degree of population ageing, and other diseases are uneven globally and could impact the population at risk of developing CAVD^[19]^. It is also possible that some of the observed variation reflects regional differences in what is recognised as CAVD and reporting systems in place. This concern might be magnified in regions where access to diagnostic services is limited.

The geographical distribution of calcific AS is heterogeneous and displays a clustering effect^[20, 21]^. Areas with the highest prevalence of CAVD tend to have the highest cause-specific CAVD mortality rates. This finding could reflect regional differences in case fatality rates driven by the availability of treatment or differential comorbidities that have a substantial impact on outcomes. Alternatively, in a recurrent theme, some of the difference might be explained by regional differences in disease attribution. Disease labelled as CAVD might be distinct in different regions and might include entities with a higher or lower case fatality rate in different regions. Although we excluded these relative differences, absolute differences in regional prevalence and cause-specific mortality were small as a proportion of the total population. This observation may suggest that these combined factors have an equally small impact on the population.

Calcific aortic valve disease (CAVD) is the most common valve heart disease in the elderly and a leading cause of aortic valve stenosis^[22]^. Because no effective pharmacological therapy exists, CAVD confers a high clinical and economic burden. The disease progression is rapid and, although initially indolent, it results in heart failure and premature death if left untreated^[23, 24]^. The current estimates support the long-standing recognition that CAVD is more common in males but refute the historical observation that CAVD is predominantly a diagnosis of youth. Recent registries report that CAVD is being diagnosed at an increasing age in older adults, an observation that is also reflected in the GBD estimates. CAVD prevalence was highest among individuals aged 90–94 years in the GBD estimates. The current estimates can confirm that physicians are increasingly labelling cardiopulmonary diseases as CAVD among older adults worldwide, and further research is needed to understand the key elements this shift in population-level driving demographics^[25]^.

This analysis of the GBD database adds to the knowledge of the epidemiology of CAVD. Geographical disparities in mortality reflect differences in valvular interventions but also many other aspects of healthcare, such as cardiovascular prevention, diagnosis of valvular disease, and follow-up. Anticipating increased burden of CAVD is particularly relevant in countries where future changes in population structure are expected to occur. Populations aging is a general trend in most countries and a consequence is at least a doubling number of patients with aortic stenosis in the next 50 years in Europe and in North America^[2, 26, 27]^. In a long-term perspective, this will result in important cumulative morbidity and mortality over time^[28]^. The current study offers comprehensive cause-specific mortality rates and the burden associated with CAVD was substantial, with a global impact of 142 000 mortality contributed by CAVD in 2021. Despite a higher prevalence in males, the burden of disease was similar for males and females, which supports the hypothesis that the disease course might be more aggressive in males. The sex paradox in CAVD—higher prevalence in males but higher case fatality rate in females—is well recognised but not fully explained, and could be related to sex hormones, health behaviours, and differences in left heart adaptation among other factors. Nevertheless, estimates of burden appear to be improving over time. Our work does not fully clarify whether these improvements are related to improved treatment, changing disease definitions, risk factor mitigation, or competing risks driving mortality towards other causes.

Estimation of CAVD burden in GBD could be an important epidemiological tool to identify emerging problems contributing to higher local or regional prevalence. This recognition, in turn, could focus research and resources to better prevent, identify, and treat CAVD in the future. This work establishes a baseline for ongoing longitudinal monitoring of CAVD prevalence against which future deviations can be compared or against which a health-care system can estimate the degree to which CAVD diagnoses might be missed.

The interpretation of CAVD mortality in the GBD is conplex due to the use of International Classification of Diseases and Injuries 10th Revision (ICD-10) codes to identify patients with CAVD. A drawback of ICD-10 is that they do not provide information on the severity of valvular stenosis or regurgitations, symptoms and consequences on left ventricle^[29]^. A number of deaths are probably related to the haemodynamic consequences of aortic stenosis, when aortic stenosis is severe and when patients are symptomatic. On the other hand, patients with an ICD-10 code of aortic stenosis or regurgitation who have non-severe aortic valve disease may have excess mortality for other reasons^[29]^. In these cases, increased cardiovascular mortality is likely related to consequences of atherosclerosis, which shares risk factors and pathophysiological pathways with CAVD^[1]^. The development of CAVD is increasingly considered to be an atherosclerosis-like process, especially in the initiation phase^[30]^. Both CAVD and atherosclerosis represent chronic inflammatory disorders, which involve initial endothelial damage and activation, lipid deposition, immune cell recruitment, inflammation, neoangiogenesis and calcification^[30–32]^. Although it attracts comparatively less attention and is less mature than mechanistic research in atherosclerosis, numerous studies have elucidated the molecular biology underlying CAVD and point to several potentially promising drug targets^[33]^. For example, a causal role for lipoprotein(a) [Lp(a)] has been demonstrated. The meta-analysis of existing evidence implies a link between elevated Lp(a) and increased incidence of aortic valve disease^[34]^, with increased mortality and risk for serious adverse outcomes associated with higher Lp(a) levels^[35]^.

Although data limitations have been acknowledged and the estimates herein might be regarded as the lower bound of the CAVD burden, it’s remarkable that, for a disease long considered among degenerative heart valve diseases until recently, there is a relatively high degree of data availability and consistency across different regions^[36–38]^. Looking ahead, maintaining unwavering attention to clear disease definitions, making progress in testing and screening procedures, and strengthening reporting systems are of critical importance. This is essential to guarantee that, when diagnosing CAVD, countries are consistently identifying the same disease, and thus coming closer to approximating the full extent of the condition’s burden. As costly CAVD treatments keep emerging and the population ages further, these aspects of health policy will grow in significance. By staying vigilant, we can ensure proper resource allocation, the alleviation of local risk factors, drugs are sustainably targeted, and prevent systemic oversights in CAVD diagnoses within health systems.

In summary, this study harnesses all accessible data, along with covariates and sophisticated models, to uncover the global epidemiology of calcific aortic valve disease (CAVD) with a finer demographic resolution than prior research. Our findings suggest that in 2021, at least 13.32 million individuals were affected by CAVD worldwide, and regrettably, 142,000 succumbed to the disease. It is evident that CAVD has a higher prevalence among males and the elderly. A more comprehensive understanding of the global epidemiological landscape enables us to strategically allocate resources and direct research efforts towards the regions, genders, and age brackets that are most in urgent need. Moreover, it may also help in spotting emerging risk factors that could potentially trigger CAVD in the days to come.

## 5. Conclusion

Over the past three decades, the Global Burden of Disease (GBD) initiative has thrived, primarily because it has met a crucial requirement in the realm of global health. We are certain that, in the future, the demand for current, pertinent, and reliable health information will persist, given the multitude of factors influencing population health. As decision-makers encounter ever more intricate choices regarding health systems and public health, their need for up-to-the-minute data, along with the translation of this information into policy-oriented speculative scenarios, will probably grow. It is our aspiration that the GBD initiative will carry on for another 30 years, playing a role in chronicling the journey of humanity in relation to health.

## 6. Limitation

This study is subject to several limitations. To begin with, the estimates in this paper are not all-encompassing. Our focus was confined to non-rheumatic calcific aortic valve disease, which were treated as a level-four disease, rather than level-three "non-rheumatic valvular heart disease." In some underdeveloped nations with relatively low healthcare standards, misdiagnosis and underdiagnosis of this condition are likely to occur. This, in turn, leads to an underestimation of the disease burden. Secondly, the data sourced from the GBD leans heavily on modelled data. GBD collaborators employ a multitude of statistical modelling techniques, especially in countries where primary data is scarce. Thirdly, the burden of CAVD is complex to define under one aspect, the disability weights definitions are still very partial as a result, reliable "measurability" of the disability is very difficult. Additionally, it’s crucial to acknowledge the time lag inherent in GBD data. Consequently, there is a dual necessity. On one hand, scales and coefficients related to patient disability require further refinement. This could potentially influence future GBD definitions of CAVD. On the other hand, more real-world studies are essential for validating the results, enabling a more precise and comprehensive evaluation.

## Data Availability

All files are available from the GBD database (Landing site http://ghdx.healthdata.org/gbd-results-tool).

https://www.healthdata.org/research-analysis/about-gbd

## Acknowledgments

The authors would like to thank the participants and researchers involved in this study; without them, this study would not have been possible.

